# Expression levels of *HLA-DRB* and *HLA-DQ* are associated with MHC Class II haplotypes in healthy individuals and rheumatoid arthritis patients

**DOI:** 10.1101/2021.01.06.20249032

**Authors:** Miranda Houtman, Anna Dzebisashvili, Espen Hesselberg, Anatoly Dubnovitsky, Genadiy Kozhukh, Lars Rönnblom, Lars Klareskog, Vivianne Malmström, Leonid Padyukov

**Affiliations:** Division of Rheumatology, Department of Medicine, Karolinska Institutet, Karolinska University Hospital, Stockholm, Sweden; Department of Medical Sciences, Rheumatology, Science for Life Laboratory, Uppsala University, Uppsala, Sweden

**Keywords:** Major histocompatibility complex, RNA-sequencing, Allelic expression, T cell, Monocyte

## Abstract

*HLA-DRB1* alleles have been associated with several autoimmune diseases. In anti-citrullinated protein antibody positive rheumatoid arthritis (ACPA-positive RA), *HLA-DRB1* shared epitope (SE) alleles are the major genetic risk factors. In order to investigate whether expression of different alleles of major histocompatibility complex (MHC) Class II genes influence functions of immune cells, we investigated transcriptomic profiles of a variety of immune cells from healthy individuals carrying different *HLA-DRB1* alleles. Sequencing libraries from peripheral blood mononuclear cells, CD4+ T cells, CD8+ T cells, and CD14+ monocytes of 32 genetically pre-selected healthy female individuals were generated, sequenced and reads were aligned to the standard reference. For the MHC region, reads were mapped to available MHC reference haplotypes and AltHapAlignR was used to estimate gene expression. Using this method, *HLA-DRB* and *HLA-DQ* were found to be differentially expressed in different immune cells of healthy individuals as well as in whole blood samples of RA patients carrying *HLA-DRB1* SE-positive versus SE-negative alleles. In contrast, no genes outside the MHC region were differentially expressed between individuals carrying *HLA-DRB1* SE-positive and SE-negative alleles. Existing methods for HLA-DR allele-specific protein expression were evaluated but were not mature enough to provide appropriate complementary information at the protein level. Altogether, our findings suggest that immune effects associated with different allelic forms of HLA-DR and HLA-DQ may be associated not only with differences in the structure of these proteins, but also with differences in their expression levels.

## 1. Introduction

Rheumatoid arthritis (RA) is a chronic inflammatory disorder affecting approximately 0.5-1% of the population worldwide [1]. Although the exact cause of RA remains unknown, a set of so-named shared epitope (SE) alleles, *HLA-DRB1*\*01 (*01:01 and *01:02), *04 (*04:01, *04:04, *04:05, and *04:08), and *10 (*10:01), have been associated with RA [2] and more specifically with anti-citrullinated protein antibody (ACPA)-positive RA [3]. These alleles share a sequence encoding five amino acids in position 70-74 of the antigen-binding groove of the HLA-DR beta chain. More recent studies have shown the importance also of amino acids in positions 11 and 13 in the HLA-DR beta chain [4]. It has been suggested that citrullinated antigens may bind preferentially to HLA-DRB1 SE sequences leading to the activation of autoreactive T-cells [5]. *HLA-DRB1* SE alleles are not the only alleles associated with ACPA-positive RA, a meta-analysis has described that *HLA-DRB1*\*13:01 alleles provide protection against ACPA-positive RA [6]. Indeed, many autoimmune diseases are associated with certain *HLA-DRB1* alleles. For example, *HLA-DRB1*\*15:01 confers the strongest risk for developing multiple sclerosis [7], but is not associated with RA. Moreover, the genetic architecture of the HLA locus is complex as allelic variants of *HLA-DRB1* involve linkage with either none or one of the paralogs (*HLA-DRB3, HLA-DRB4* or *HLA-DRB5*). Therefore, *HLA-DRB1* is expressed in cells with all *HLA-DRB1* haplotypes, whereas expression of the paralogs is haplotype-specific (Figure 1).

**Figure 1.**
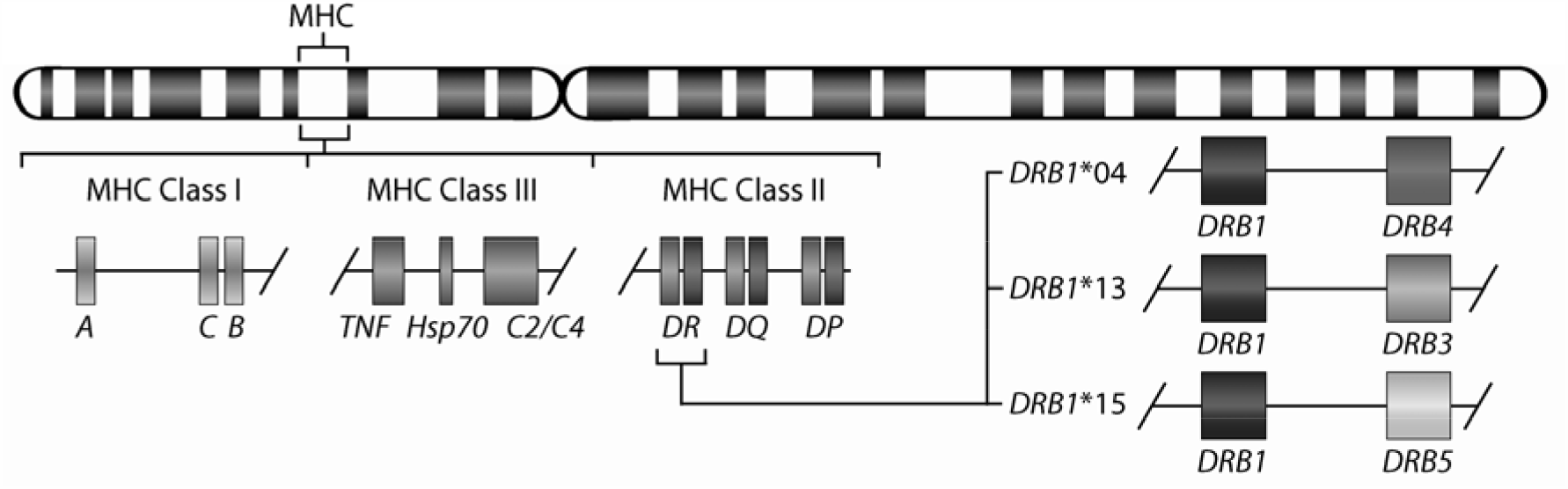
Gene map of the major histocompatibility complex (MHC) region. The MHC region on the short arm of human chromosome 6, contains the HLA-DR (HLA-DRA and HLA-DRB) molecules. The allelic variants of *HLA-DRB1* (*DRB1*\*04, *13, and *15) are linked with only one of the genes *HLA-DRB4*, or *HLA-DRB3*, or *HLA-DRB5*.

The genetic association of *HLA-DRB1* to RA is proposed to at least partly reflect a favoured binding of citrullinated peptides to the HLA binding groove. However, the precise molecular mechanisms by which *HLA-DRB1* SE alleles predispose to ACPA-positive RA are currently unclear. It has previously been suggested that differences in expression of genes in the major histocompatibility complex (MHC) region between different HLA-DRB1 haplotypes could contribute to disease [8-10], but this has not yet been studied in RA patients and for specific cell types. In this study, we aimed to identify differentially expressed genes in peripheral blood mononuclear cells (PBMCs) and isolated CD4+ T cells, CD8+ T cells, and CD14+ monocytes from PBMCs of healthy individuals with *HLA-DRB1* SE alleles compared to healthy individuals not carrying these alleles, which will be referred to as SE negative alleles. In addition, the total expression of the MHC Class II molecules in whole blood samples was investigated in both healthy individuals and in RA patients. We used RNA-seq data with a pipeline (AltHapAlignR [8]) that aligns reads to available MHC reference haplotypes for more accurate analysis of gene expression in the MHC region.

## 2. Materials and Methods

### 2.1. Blood donors

Blood samples from 32 healthy previously genotyped donors (females between 51 and 80 years of age) were provided by the Uppsala Bioresource (Uppsala University Hospital, Uppsala, Sweden). This project was undertaken with ethical approval of the regional ethical review board in Uppsala and all healthy donors gave informed consent according to the declaration of Helsinki. The samples were selected by positivity for certain *HLA-DRB1* alleles (*HLA-DRB1\**04, *HLA-DRB1*\*13:01, and *HLA-DRB1*\*15:01). The main characteristics of the participants are presented in Table 1.

**Table 1.** Characteristics of study participants.

| **Sample ID** | **Sex** | **Age range** | **HLA-DRB1 alleles** | **HLA-DRB1 SE** | **PBMCs** | **CD4+ T cells** | **CD8+ T cells** | **CD14+ monocytes** |
| --- | --- | --- | --- | --- | --- | --- | --- | --- |
| 1 | F | 51-60 | *03:01/*04:01 | Positive | x | x | x | NA |
| 2 | F | 51-60 | *03:01/*04:01 | Positive | x | NA | NA | NA |
| 3 | F | 51-60 | *03:01/*15:01 | Negative | x | NA | x | NA |
| 4 | F | 61-70 | *03:01/*15:01 | Negative | NA | NA | x | NA |
| 5 | F | 51-60 | *01:01/*04:01 | Double positive | x | x | x | NA |
| 6 | F | 61-70 | *03:01/*13:01 | Negative | x | x | NA | NA |
| 7 | F | 61-70 | *04:01/*07:01 | Positive | x | x | x | NA |
| 8 | F | 51-60 | *04:01/*13:03 | Positive | NA | x | x | NA |
| 9 | F | 61-70 | *04:01/*07:01 | Positive | x | x | NA | x |
| 10 | F | 51-60 | *03:01/*04:01 | Positive | x | x | NA | x |
| 11 | F | 61-70 | *03:01/*15:01 | Negative | x | x | NA | NA |
| 12 | F | 61-70 | *04:01/*14:01 | Positive | x | x | x | NA |
| 13 | F | 61-70 | *03:01/*13:01 | Negative | NA | x | NA | NA |
| 14 | F | 61-70 | *03:01/*15:01 | Negative | x | x | NA | NA |
| 15 | F | 61-70 | *01:01/*04:01 | Double positive | x | x | x | x |
| 16 | F | 51-60 | *03:01/*04:01 | Positive | x | x | x | NA |
| 17 | F | 61-70 | *03:01/*15:01 | Negative | x | x | NA | x |
| 18 | F | 61-70 | *03:01/*13:01 | Negative | x | x | x | NA |
| 19 | F | 61-70 | *03:01/*04:01 | Positive | x | x | NA | NA |
| 20 | F | 51-60 | *03:01/*13:01 | Negative | x | x | x | NA |
| 21 | F | 71-80 | *03:01/*13:01 | Negative | x | x | NA | NA |
| 22 | F | 51-60 | *04:01/*04:04 | Double positive | x | NA | x | NA |
| 23 | F | 51-60 | *03:01/*13:01 | Negative | x | x | x | NA |
| 24 | F | 61-70 | *04:01/*14:01 | Positive | x | NA | NA | x |
| 25 | F | 61-70 | *04:01/*09:01 | Positive | x | x | x | NA |
| 26 | F | 61-70 | *03:01/*04:01 | Positive | x | NA | x | NA |
| 27 | F | 71-80 | *01:01/*04:01 | Double positive | x | x | x | x |
| 28 | F | 61-70 | *03:01/*13:01 | Negative | x | x | x | x |
| 29 | F | 51-60 | *04:08/*07:01 | Positive | x | x | x | NA |
| 30 | F | 61-70 | *03:01/*15:01 | Negative | x | x | x | x |
| 31 | F | 61-70 | *03:01/*15:01 | Negative | x | x | x | x |
| 32 | F | 51-60 | *01:01/*04:01 | Double positive | x | x | x | x |

Abbreviations: F – female, NA – not available, PBMCs – peripheral blood mononuclear cells, SE – shared epitope, x – included in analyses.

### 2.2. Blood sampling and cell separation

Buffy coats were processed by density gradient centrifugation using Ficoll (GE Healthcare Bio-Sciences AB, Uppsala, Sweden) and PBMCs were subsequently recovered. CD4+, CD8+, and CD14+ cells were isolated from the PBMCs *via* positive selection using CD4, CD8 or CD14 Microbeads (Miltenyi Biotec Norden AB, Lund, Sweden) on the autoMACS® Pro Separator (Miltenyi Biotec Norden AB).

### 2.3. RNA sequencing

Total RNA was extracted with the RNeasy Mini kit (Qiagen AB, Sollentuna, Sweden) according to manufacturer’s instructions. Samples were treated with DNase (Qiagen) for 20 min at room temperature to avoid contamination with genomic DNA. The quality of each RNA sample was assessed using the Agilent Bioanalyzer 2100 and RNA 6000 Nano Chips (Agilent Technologies Sweden AB, Kista, Sweden). The RNA integrity number (RIN) ranged between 3.4 and 9.5 (median of 7.9). The RNA was fragmented and prepared into sequencing libraries using the Illumina TruSeq stranded total RNA sample preparation kit with ribosomal depletion using RiboZero (2 x 125 bp) and sequenced on an Illumina HiSeq 2500 sequencer (SNP&SEQ Technology Platform, Uppsala, Sweden). Between 24.5 and 60.5 (median of 41) million reads were produced per sample. Raw read quality was evaluated using FastQC. Pre-filtering on quality of reads using cutadapt (version 1.9.1) was applied (-q 30 -a AGATCGGAAGAGCACACGTCTGAACTCCAGTCAC -A AGATCGGAAGAGCGTCGTGTAGGGAAAGAGTGTAGATCTCGGTGGTCGCCGTATCATT -m 40). Filtered reads were aligned to the hg38 assembly, containing the MHC reference haplotype PGF (GENCODE release 28 (GRCh38.p12)), using STAR in two-pass mode (version 2.5.4b) [11] with default settings. STAR was also used to obtain the gene counts. All sequencing data generated in this study are available at NCBI Gene Expression Omnibus accession number GSE163605.

### 2.4. Whole blood RNA sequencing data of healthy individuals and RA patients

FASTQ files for 439 whole blood samples were downloaded from the GTEx project [12]. For RA patient samples, 158 FASTQ files from the EIRA/RECOMBINE project were used. Seq2HLA [13] was used to impute classical HLA alleles from RNA-seq data. Samples with *HLA-DRB1*\*03, *04, *07 and *15 alleles were selected according to available MHC reference haplotypes. Mapping to the human genome, MHC region and differential gene expression analysis were performed as described above.

### 2.5. Mapping to the MHC region

Counts of genes in the MHC region (chr6: 28,500,000-33,500,000) were replaced with counts obtained from AltHapAlignR [8]. In short, unmapped reads and reads mapped to the MHC region (using MHC reference haplotype PGF) were extracted and realigned to all available MHC reference haplotypes (APD, COX, DBB, MANN, MCF, QBL, SSTO) independently using STAR in two-pass mode. Reads mapped to multiple regions, with mapping quality less than 20, and duplicate reads were removed.

### 2.6. Differential gene expression analysis

Raw expression counts obtained from STAR (non-MHC genes) and AltHapAlignR (MHC genes) were adjusted for library size using the R package DESeq2 (version 1.26.0) [14]. Pre-filtering of low count genes was performed to keep only genes that had at least 50 counts in total over all samples. The counts were regularized-logarithm transformed for principal component analysis (PCA). For each specific cell subset, RIN score was highly correlated with principal component 1 and used as covariate as binary category (< 7 and ≥ 7) in the analyses. The default DESeq2 options were used, including log fold change shrinkage using apeglm [15] and independent hypothesis weighting [16]. *P*-values were obtained with the Wald test. Differences in gene expression with Benjamini-Hochberg adjusted *P*-value < 0.05 and fold change (log2) > 1 were considered significant.

### 2.7. HLA-typing and imputation

HLA-DRB1 genotypes were initially imputed from Immunochip data by SNP2HLA [17] and later HLA low resolution typing was performed for validation. Additionally, DR4 subtyping was performed for *HLA-DRB1*\*04 positive individuals. *HLA*-typing was performed by sequence-specific primer polymerase chain reaction assay (DR low-resolution kit and DR4 kit; Olerup SSP, Stockholm, Sweden) and analyzed by agarose gel electrophoresis [18]. An interpretation table was used to determine the specific genotype according to the manufacturer’s instructions. In addition, seq2HLA [13] was used to impute classical HLA alleles from RNA-seq data.

### 2.7. Quantitative real-time PCR

RNA of the PBMC, CD4+, CD8+, and CD14+ cell subset samples was converted into cDNA using iScript™ Reverse Transcription Supermix (Bio-Rad, Solna, Sweden). Quantitative real-time PCR (qPCR) was carried out using SsoAdvanced™ Universal SYBR Green Supermix (Bio-Rad) and primers detecting *HLA-DQA* (forward primer 5’- CAACATCACATGGCTGAGCA-3’ and reverse primer 5’- TGCTCCACCTTGCAGTCATAA-3’), *HLA-DQB* (forward primer 5’- TCTCCCCATCCAGGACAGAG-3’ and reverse primer 5’- TTCCGAAACCACCGGACTTT-3’), and *HLA-DRA* (forward primer 5’- CCTGTCACCACAGGAGTGTC-3’ and reverse primer 5’- TCCACCCTGCAGTCGTAAAC-3’) on a CFX384 Touch™ system (Bio-Rad) with the following protocol: 95°C for 30 sec, followed by 40 cycles of 95°C for 10 sec and 60°C for 30 sec. The endogenous controls *ACTIN* (forward primer 5’- GGACTTCGAGCAAGAGATGG-3’ and reverse primer 5’- AGCACTGTGTTGGCGTACAG-3’), *UBE2D2* (forward primer 5’-TGCCTGAGATTGCTCGGATCT-3’ and reverse primer 5’- TCGCATACTTCTGAGTCCATTCC-3’), and *ZNF592* (forward primer 5’-GTAAAGGAGAATTGCCTGCA-3’ and reverse primer 5’- GAATGCACATTTGTGGAAAA-3’). Data was analyzed with the SDS 2.4 software of Applied Biosystems before applying the ΔΔCt method [19].

### 2.9 Cell type enrichment analysis

The xCell tool [20] was used to estimate cellular heterogeneity in the PBMC, CD4+, CD8+, and CD14+ cell subsets from RNA sequencing data. xCell uses the expression levels ranking (Transcripts Per Million (TPM)) and these were obtained using Salmon (version 0.8.2) [21].

### 2.10. Expression and purification of HLA-DRB recombinant proteins

The extracellular domains of the HLA-DRB1*03:01, 04:01, 15:01 and HLA-DRB5*01:01 beta chains with a basic leucine zipper were cloned into pET28a vector and expressed in *E*.*coli* BL21 (DE3) STAR cells (Novagen). Inclusion bodies from *E*.*coli* cells were dissolved in 8 M Urea, 50 mM TRIS-HCl buffer pH 8. Proteins were purified using a combination of anion-exchange chromatography on MonoQ 5/50 column (GE Healthcare) followed by size-exclusion step using Superdex 200 10/300 GL column (GE Healthcare). Protein concentrations were determined based on absorption at 280nm using theoretical extinction coefficients calculated by ProtParam server (https://web.expasy.org/protparam/) from amino acid sequences.

### 2.11. Protein extraction and western blotting

PBMC samples that had previously been collected and frozen in liquid nitrogen were cultured in RPMI-1640 medium (Sigma-Aldrich, Stockholm, Sweden) supplemented with 10% fetal bovine serum (Sigma-Aldrich), L-glutamine (Gibco, Stockholm, Sweden) and penicillin/streptomycin (Gibco) in a 37°C incubator with 5% CO_2_. After 40 hours, cell lysates were prepared using RIPA buffer (ThermoFisher Scientific, Stockholm, Sweden) including 3× Halt™ Protease Inhibitor Cocktail (ThermoFisher Scientific). Protein concentrations were quantified using the Pierce™ BCA Protein Assay (ThermoFisher Scientific). Cell lysates and recombinant proteins were mixed with NuPAGE™ LDS Sample Buffer and NuPAGE™ Sample Reducing Agent, heated at 70°C for 10 min and loaded onto NuPAGE™ 4-12% gradient Bis-Tris gels (ThermoFisher Scientific). Proteins were transferred to a membrane using the iBlot® Gel Transfer Device (ThermoFisher Scientific). Membrane was blocked with 5% non-fat milk in TBST at room temperature for 1 hour. Primary antibodies were diluted in 5% non-fat milk in TBST (ab133578 – rabbit monoclonal to HLA Class II DRB1 (1:5000 (Abcam, Amsterdam, the Netherlands)), DA2 – mouse monoclonal to HLA-DRB (1:1000 (Santa Cruz Biotechnology, Heidelberg, Germany)), ab202343 – rabbit polyclonal to HLA Class II DRB1 (1:4000 (Abcam)), ab98108 – rabbit polyclonal to HLA Class II DRB1 (1:2000 (Abcam)), ab168534 – rabbit polyclonal to HLA Class II DRB1 (1:2000 (Abcam)), and E1E9V – rabbit monoclonal to vinculin (1:1000 (Cell Signaling Technology, Solna, Sweden) and incubated overnight at 4°C. Secondary antibodies were diluted in 5% non-fat milk in TBST (polyclonal goat anti-mouse HRP (1:5000 (Agilent, Kista, Sweden)) and polyclonal goat anti-rabbit HRP (1:2500 (Santa Cruz Biotechnology)) and incubated at room temperature for 1 hour. Membranes were incubated with SuperSignal™ West Pico PLUS Chemiluminescent Substrate (ThermoFisher Scientific) and chemiluminescence was captured using the ChemiDoc™ MP Imaging System (Bio-Rad). Image Lab 6.0 (Bio-Rad) was used to determine the intensities of identified bands.

## 3. Results

### 3.1. Differentially expressed genes in PBMCs of *HLA-DRB1* SE-positive versus SE-negative healthy individuals

To identify differentially expressed genes in PBMCs of healthy individuals with and without *HLA-DRB1* SE alleles, we conducted RNA-seq on 29 PBMC samples and aligned reads to available MHC reference haplotypes using AltHapAlignR [8]. We identified five MHC Class II genes (*HLA-DRB4, HLA-DQA2, HLA-DRB1, HLA-DQA1*, and *HLA-DQB1*) that were differentially expressed to a great degree in PBMCs of *HLA-DRB1* SE-positive versus SE-negative individuals (Figure 2A and Table S1). Of these differentially expressed genes, *HLA-DRB4, HLA-DQA2, HLA-DRB1*, and *HLA-DQA1* were higher expressed in *HLA-DRB1* SE-positive individuals, whereas *HLA-DQB1* was lower expressed in *HLA-DRB1* SE-positive individuals, in comparison to *HLA-DRB1* SE-negative individuals. The expression differences of *HLA-DQA* and *HLA-DQB* depending on *HLA-DRB1* alleles could be confirmed by qPCR (Figure S1 and S2). We identified no significantly differentially expressed genes outside the MHC region in PBMCs of *HLA-DRB1* SE-positive versus SE-negative healthy individuals with an adjusted *P*-value < 0.05 and a fold change (log2) > 1 (Table S1).

**Figure 2.**
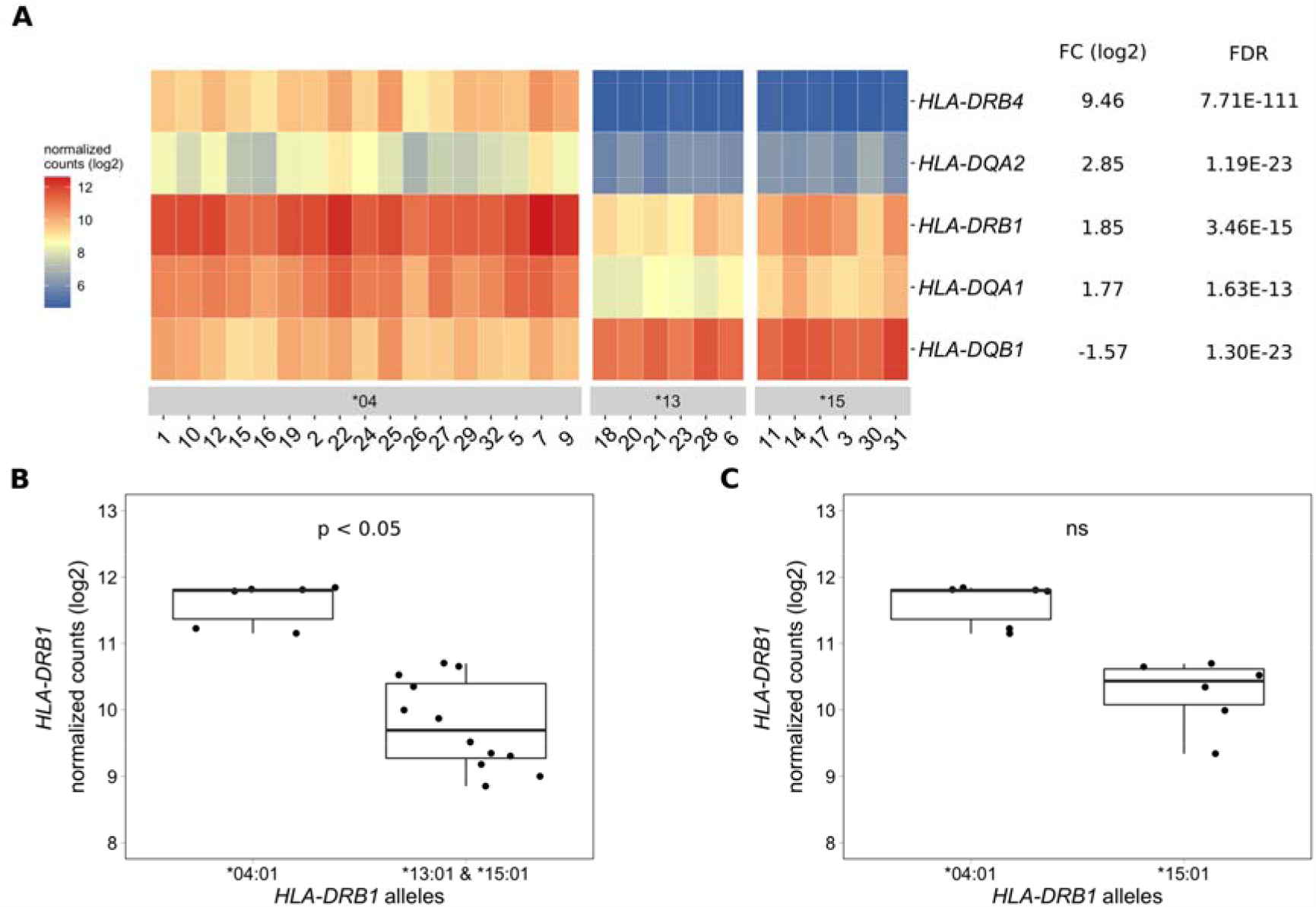
Differentially expressed MHC Class II genes in PBMCs of *HLA-DRB1* SE-positive versus SE-negative healthy individuals. A) Heat map of differentially expressed genes (log transformed normalized gene counts) in PBMCs of *HLA-DRB1* SE-positive [*04 (*n* = 17)] versus SE-negative [*03:01/*13:01 and *03:01/*15:01 (*n* = 12)] individuals. Genes shown in red have higher gene counts and those shown in blue have lower gene counts. B) *HLA-DRB1* gene expression in PBMCs of *HLA-DRB1* SE-positive [*03:01/*04:01 (*n* = 6)] and SE-negative [*03:01/*13:01 and *03:01/*15:01 (*n* = 12)] individuals. C) *HLA-DRB1* gene expression in PBMCs of *HLA-DRB1* SE-positive [*03:01/*04:01 (*n* = 6)] and SE-negative [*03:01/*15:01 (*n* = 6)] individuals. FC: fold change, ns: non-significant adjusted *P*-value.

To reduce the effect of the second allele in the *HLA-DRB1* SE-positive group, we performed differential gene expression analysis on PBMC samples with only *HLA-DRB1*\*03:01 as second allele. We found that the five MHC Class II genes (*HLA-DRB4, HLA-DQA2, HLA-DRB1, HLA-DQA1*, and *HLA-DQB1*) were differentially expressed in PBMCs of *HLA-DRB1* SE-positive versus SE-negative individuals carrying one *HLA-DRB1*\*03:01 allele (adjusted *P*-value < 0.05 and a fold change (log2) > 1 (Table S1 and Figure 2B)).

As the *HLA-DRB1* sequence appeared to be absent in the MHC reference haplotype APD (*HLA-DRB1*\*13:01), we also performed differential gene expression analysis on PBMC samples of individuals with the alleles *HLA-DRB1*\*03:01/*04:01 versus *03:01/*15:01. Four MHC Class II genes (*HLA-DRB5, HLA-DRB4, HLA-DQA2*, and *HLA-DQB1*) were differentially expressed in PBMCs of healthy individuals carrying *HLA-DRB1*\*03:01/*04:01 compared to *03:01/*15:01 (adjusted *P*-value < 0.05 and a fold change (log2) > 1 (Table S1)). In addition, there is a trend towards *HLA-DRB1* being higher expressed in PBMCs of individuals carrying *HLA-DRB1*\*03:01/*04:01 compared to *HLA-DRB1*\*03:01/*15:01, although no statistically significant difference was observed (adjusted *P*-value of 0.068 and fold change (log2) of 0.904 (Figure 2C)).

### 3.2. Differentially expressed genes in CD4+ and CD8+ T cells of *HLA-DRB1* SE-positive versus SE-negative healthy individuals

As PBMCs are a heterogeneous mixture of immune cell types, we isolated CD4+ T cells, CD8+ T cells, and CD14+ monocytes from the same healthy individuals *via* positive selection using microbeads and performed differential gene expression analyses. We found five MHC Class II genes (*HLA-DRB4, HLA-DQA2, HLA-DRB1, HLA-DQA1*, and *HLA-DQB1*) that were differentially expressed to a great degree in CD4+ T cells of *HLA-DRB1* SE-positive versus SE-negative individuals (Figure 3A, 3C and Table S1). Again, we identified no differentially expressed genes outside the MHC region in CD4+ T cells of *HLA-DRB1* SE-positive versus SE-negative individuals (adjusted *P*-value < 0.05 and a fold change (log2) > 1 (Table S1)). In CD8+ T cells, we found five MHC Class II genes (*HLA-DRB4, HLA-DRB1, HLA-DQB1, HLA-DQA2*, and *HLA-DQA1*) and two genes outside the MHC region (*RPL10P6* at chromosome 2q35 and *ADRB1* at chromosome 10q25) to be differentially expressed between *HLA-DRB1* SE-positive and SE-negative individuals (Figure 3B and Table S1). The expression differences of *HLA-DQA* and *HLA-DQB* depending on *HLA-DRB1* alleles could be confirmed by qPCR (Figure S1 and S2). In addition, there is a trend towards *HLA-DRB1* being higher expressed in CD8+ T cells of individuals carrying *HLA-DRB1*\*03:01/*04:01 compared to *HLA-DRB1*\*03:01/*15:01, although no statistically significant difference was observed (adjusted *P*-value of 0.538 and fold change (log2) of 0.811 (Figure 3D)).

**Figure 3.**
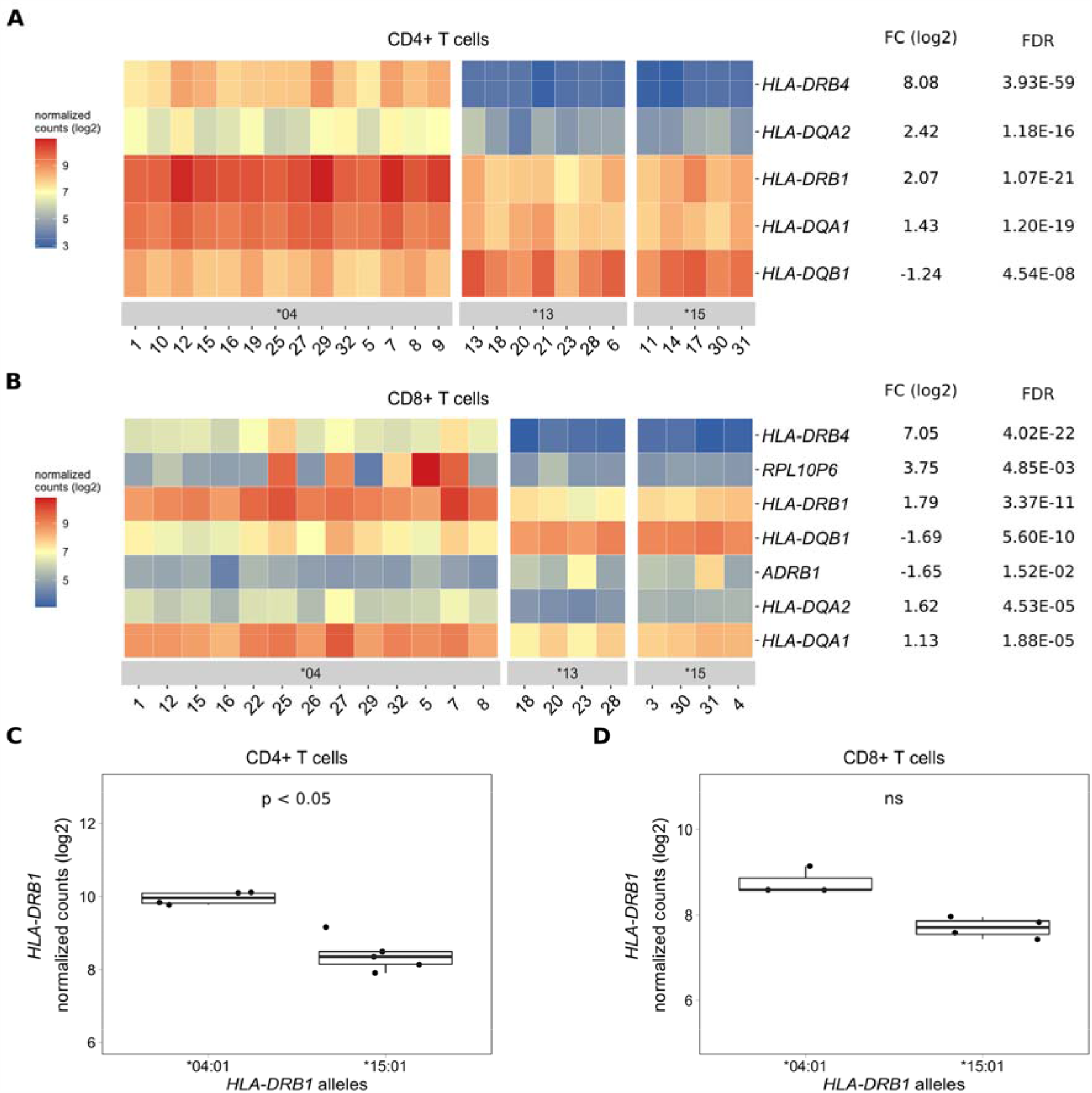
Differentially expressed genes in CD4+ and CD8+ T cells of *HLA-DRB1* SE-positive versus SE-negative healthy individuals. A) Heat map of differentially expressed genes (log transformed normalized gene counts) in CD4+ T cells of *HLA-DRB1* SE-positive [*04 (*n* = 14)] versus SE-negative [*03:01/*13:01 and *03:01/*15:01 (*n* = 12)] individuals. B) Heatmap of differentially expressed genes (log transformed normalized gene counts) in CD8+ T cells of *HLA-DRB1* SE-positive [*04 (*n* = 13)] versus SE-negative [*03:01/*13:01 and *03:01/*15:01 (*n* = 8)] individuals. In both heatmaps, genes shown in red have higher gene counts and those shown in blue have lower gene counts. C) *HLA-DRB1* gene expression in CD4+ T cells of *HLA-DRB1* SE-positive [*03:01/*04:01 (*n* = 4)] and SE-negative [*03:01/*15:01 (*n* = 5)] individuals. D) *HLA-DRB1* gene expression in CD8+ T cells of *HLA-DRB1* SE-positive [*03:01/*04:01 (*n* = 3)] and SE-negative [*03:01/*15:01 (*n* = 4)] individuals. FC: fold change, ns: non-significant adjusted *P*-value.

### 3.3. Differentially expressed genes in CD14+ monocytes of *HLA-DRB1* SE-positive versus SE-negative healthy individuals

In CD14+ monocytes, we identified five genes that were differentially expressed between *HLA-DRB1* SE-positive and SE-negative healthy individuals (adjusted *P*-value < 0.05 and a fold change (log2) > 1 (Figure 4A and Table S1)). Of these five differentially expressed genes, three were MHC Class II genes (*HLA-DRB4, HLA-DQA2*, and *HLA-DQA1*), one was a MHC Class I gene (*HLA-A*), and one was a non-MHC gene (*TENT4B* at chromosome 16q12). *HLA-DRB4, HLA-DQA2, HLA-DQA1*, and *HLA-A* were higher expressed in *HLA-DRB1* SE-positive individuals, whereas *TENT4B* was lower expressed in *HLA-DRB1* SE-positive individuals, in comparison to *HLA-DRB1* SE-negative individuals. In addition, there is a trend towards *HLA-DRB1* being higher expressed in CD14+ monocytes of individuals carrying *HLA-DRB1*\*03:01/*04:01 compared to *HLA-DRB1*\*03:01/*15:01 (adjusted *P*-value of 0.122 and fold change (log2) of 1.658 (Figure 4B)). The expression differences of *HLA-DQA* and *HLA-DQB* depending on *HLA-DRB1* alleles could be confirmed by qPCR (Figure S1 and S2).

**Figure 4.**
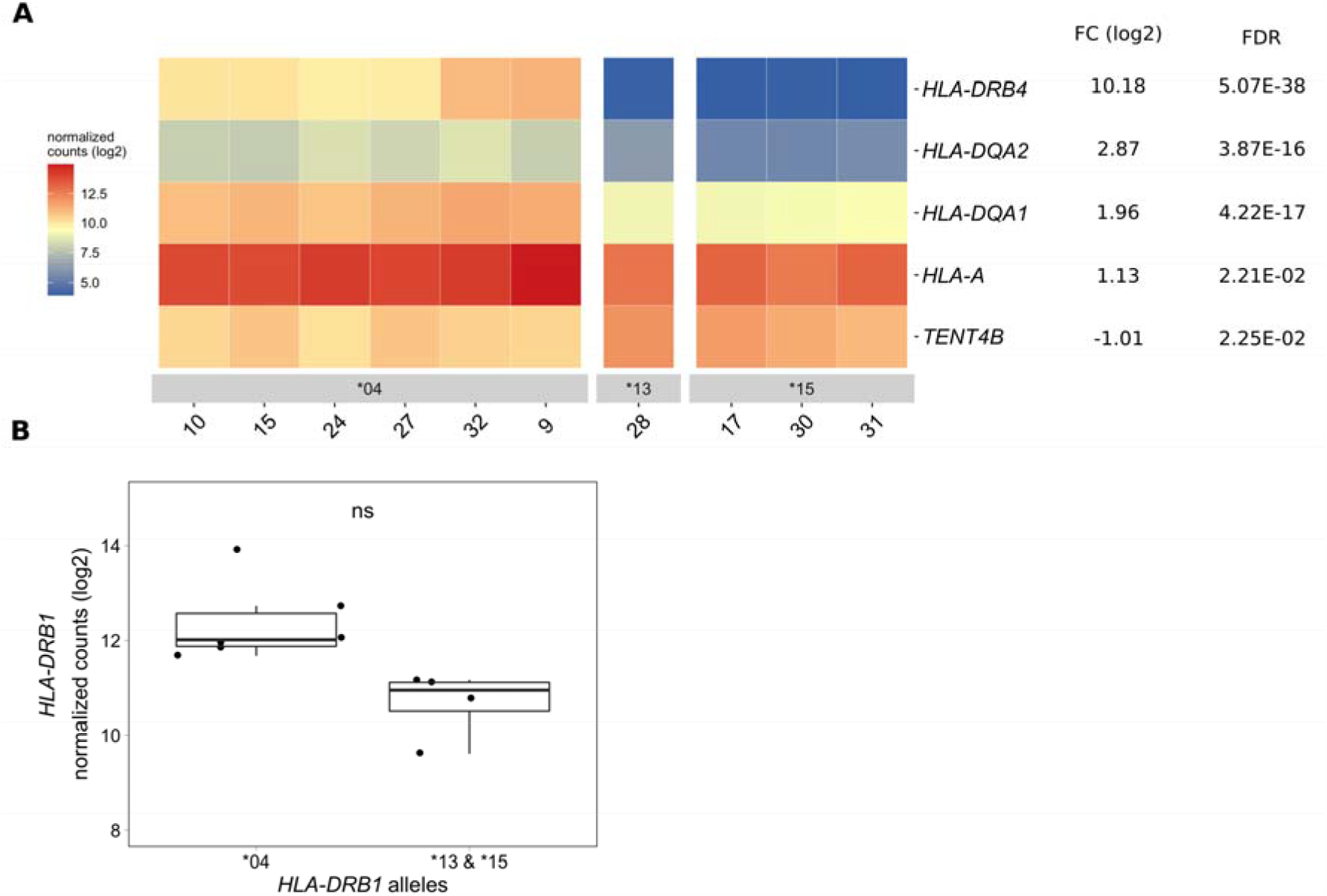
Differentially expressed genes in CD14+ monocytes of *HLA-DRB1* SE-positive versus SE-negative healthy individuals. **A)** Heat map of differentially expressed genes (log transformed normalized gene counts) in CD14+ monocytes of *HLA-DRB1* SE-positive [*04 (*n* = 6)] and SE-negative [*03:01/*13:01 and *03:01/*15:01 (*n* = 4)] individuals. Genes shown in red have higher gene counts and those shown in blue have lower gene counts for the contrast SE+ vs. SE-. **B)** *HLA-DRB1* gene expression in CD14+ monocytes of *HLA-DRB1* SE-positive [*04 (*n* = 6)] and SE-negative [*03:01/*13:01 and *03:01/*15:01 (*n* = 4)] individuals. FC: fold change, ns: non-significant adjusted *P*-value.

### 3.4. *HLA-DRB1* is differentially expressed in whole blood samples of healthy individuals carrying different *HLA-DRB1* alleles

To explore if this difference in *HLA-DRB1* gene expression could also be seen in whole blood samples of healthy individuals, we used RNA-seq data from 439 whole blood samples from the GTEx project [12]. Classical HLA alleles were imputed from RNA-seq data and samples with *HLA-DRB1*\*03, *04, *07 and *15 alleles were selected for mapping reads to the MHC region to avoid alignment biases. *HLA-DRB1* is significantly higher expressed in *HLA-DRB1* SE-positive individuals (carrying at least one *HLA-DRB1*\*04 allele) compared to *HLA-DRB1* SE-negative individuals (carrying no *HLA-DRB1*\*04 alleles (Figure 5A)). In addition, we examined the expression of *HLA-DRB1* in individuals carrying different *HLA-DRB1* alleles and determined that the expression of *HLA-DRB1* is strongly dependent on the presence of different *HLA-DRB1* alleles (Figure 5B). Within whole blood samples from the GTEx project, there is a trend towards higher expression levels of *HLA-DRB1* in *HLA-DRB1*\*04 and *07 alleles compared to *HLA-DRB1*\*03 and *15 alleles.

**Figure 5.**
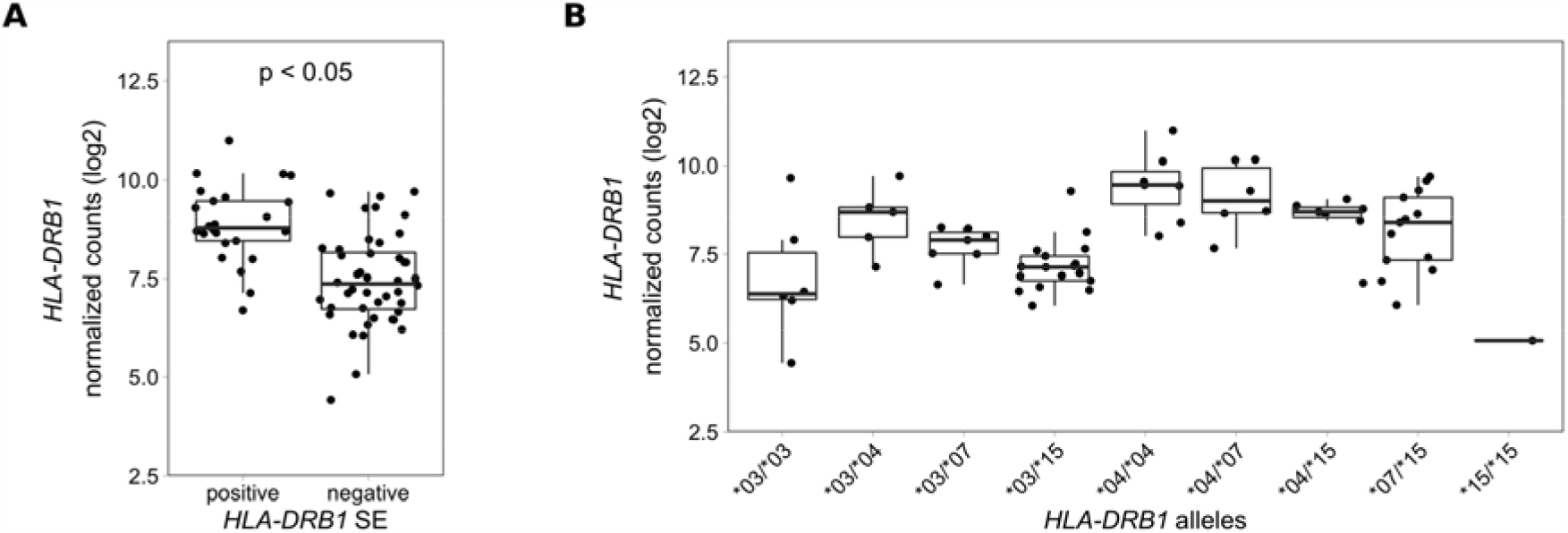
*HLA-DRB1* expression levels in whole blood samples of healthy individuals of the GTEx project. **A)** *HLA-DRB1* gene expression in whole blood samples of *HLA-DRB1* SE-positive [*04 (*n* = 25)] and SE-negative [*03, *07, and *15 (*n* = 44)] individuals. **B)** *HLA-DRB1* gene expression in whole blood samples of individuals carrying different combinations of *HLA-DRB1* alleles [*03/*03 (*n* = 6), *03/*04 (*n* = 5), *03/*07 (*n* = 7), *03/*15 (*n* = 17), *04/*04 (*n* = 7), *04/*07 (*n* = 6), *04/*15 (*n* = 7), *07/*15 (*n* = 13), and *15/*15 (*n* = 1)]. *: Adjusted *P*-value (FDR) < 0.05.

### 3.5. *HLA-DRB1* is differentially expressed in whole blood samples of RA patients carrying different *HLA-DRB1* alleles

To analyze *HLA-DRB1* expression levels in RA patients carrying different *HLA-DRB1* alleles, we used RNA-seq data from 158 whole blood samples from the EIRA/RECOMBINE project. Classical HLA alleles were imputed from RNA-seq data and samples with *HLA-DRB1*\*03, *04, *07 and *15 alleles were selected for mapping reads to the MHC region. Also in these samples, *HLA-DRB1* is significantly higher expressed in *HLA-DRB1* SE-positive RA patients (carrying at least one *HLA-DRB1*\*04 allele) compared to *HLA-DRB1* SE-negative RA patients (carrying no *HLA-DRB1*\*04 alleles) (Figure 6A). In addition, we examined the expression of *HLA-DRB1* in RA patients carrying different *HLA-DRB1* alleles and showed that the expression of *HLA-DRB1* is strongly associated with *HLA-DRB1* alleles (Figure 6B). In whole blood samples of RA patients, there is a trend towards higher expression levels of *HLA-DRB1* in *HLA-DRB1*\*04 and *07 alleles compared to *HLA-DRB1*\*03 and *15 alleles.

**Figure 6.**
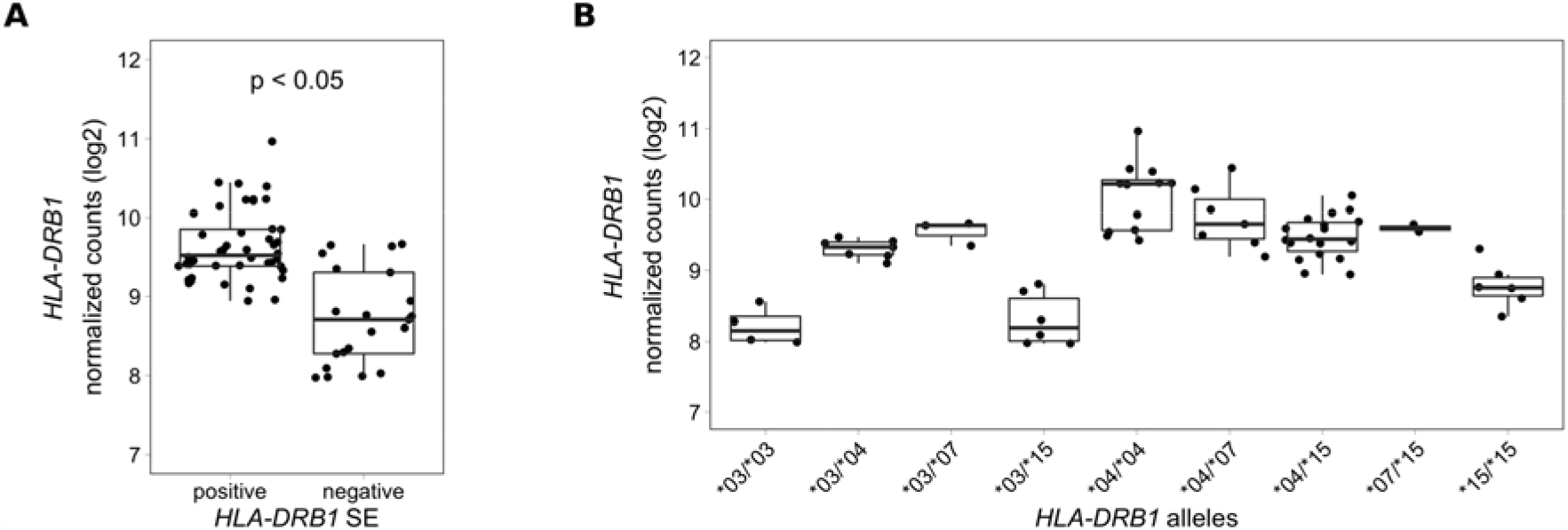
*HLA-DRB1* expression levels in whole blood samples of RA patients. **A)** *HLA-DRB1* gene expression in whole blood samples of *HLA-DRB1* SE-positive [*04 (*n* = 44)] and SE-negative [*03, *07, and *15 (*n* = 21)] RA patients. **B)** *HLA-DRB1* gene expression in whole blood samples of RA patients carrying different combinations of *HLA-DRB1* alleles [*03/*03 (*n* = 4), *03/*04 (*n* = 7), *03/*07 (*n* = 3), *03/*15 (*n* = 6), *04/*04 (*n* = 12), *04/*07 (*n* = 7), *04/*15 (*n* = 18), *07/*15 (*n* = 2), and *15/*15 (*n* = 6)]. *: Adjusted *P*-value (FDR) < 0.05.

### 3.6. HLA-DRB1 protein expression in PBMCs of *HLA-DRB1* SE-positive and SE-negative healthy individuals

To examine whether the described differences in gene expression level can be found also for expression levels of proteins, we investigated the PBMC samples of *HLA-DRB1* SE-positive and SE-negative healthy individuals by western blotting. However, an inherent problem in this approach is that antibodies may display different binding affinities to the varying HLA-DRB alleles. We tried to handle this problem by using western blot of defined amounts of recombinant HLA-DRB1*03:01, HLA-DRB1*04:01, HLA-DRB1*15:01, and HLA-DRB5*01:01 proteins as references. The binding of the tested HLA-DRB antibodies to the recombinant proteins was, however, highly heterogeneous (Figure 7A). The antibodies ab133578 and DA2 bound to all HLA-DRB1 recombinant proteins but the signal was stronger for HLA-DRB1*15:01 than for HLA-DRB1*03:01 and HLA-DRB1*04:01. The other antibodies (ab202343, ab98108, and ab168534) did not bind to HLA-DRB1*04:01 recombinant protein according to our experimental data. In addition, the antibodies ab133578 and ab202343 did bind both to HLA-DRB1 and to HLA-DRB5*01:01, thereby representing too broad specificity to enable analysis of allele specific protein expression for the HLA-DRB family of genes (Figure 7A).

**Figure 7.**
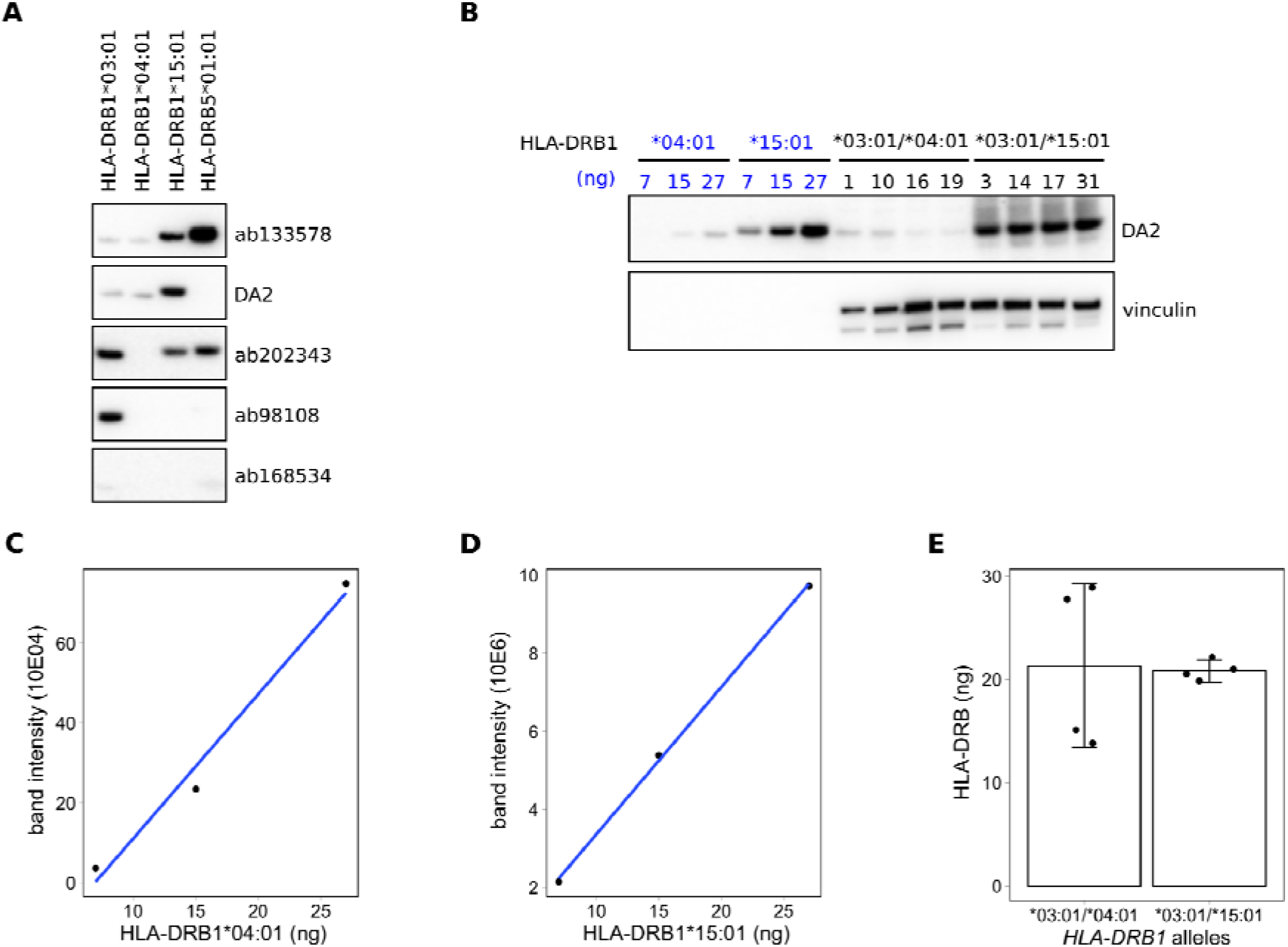
HLA-DRB protein expression in PBMCs of *HLA-DRB1* SE-positive and SE-negative healthy individuals. **A)** Representative western blots showing differential binding of different HLA-DRB antibodies (ab133578, DA2, ab202343, ab98108, and ab168534) to the recombinant proteins HLA-DRB1*03:01, HLA-DRB1*04:01, HLA-DRB1*15:01, and HLA-DRB5*01:01. **B)** Representative western blot of determining HLA-DRB levels using the antibody DA2 in PBMC samples of individuals carrying *HLA-DRB1*\*03:01/*04:01 versus *HLA-DRB1*\*03:01/*15:01 alleles. Vinculin was used as a protein loading control. **C)** Standard curve of the recombinant protein HLA-DRB1*04:01 detected by the antibody DA2. **D)** Standard curve of the recombinant protein HLA-DRB1*15:01 detected by the antibody DA2. **E)** Interpolated HLA-DRB protein levels in individuals carrying *HLA-DRB1*\*03:01/*04:01 and *HLA-DRB1*\*03:01/15:01 alleles.

In order to try to overcome these problems, we prepared standard curves of the recombinant protein HLA-DRB1*04:01 (Figure 7B and 7C) and HLA-DRB1*15:01 (Figure 7B and 7D) to enable quantification of the amount of HLA-DRB in protein extracts from PBMCs of healthy individuals carrying *HLA-DRB1*\*03:01/04:01 (*n*=4) and *HLA-DRB1*\*03:01/*15:01 (*n* = 4) alleles (Figure 7E). However, the expression of HLA-DRB was too variable in *HLA-DRB1*\*03:01/*04:01 samples, probably due to weak binding of DA2 to HLA-DRB1*04:01, to draw a firm conclusion from these experiments about HLA-DRB protein levels.

## 4. Discussion

The major finding of our study is the identification of relatively high differences in gene expression levels for several MHC Class II genes in immune cells of healthy individuals depending on MHC haplotypes, which are known as genetic risk factors for autoimmune diseases. More specifically, we found that the gene for HLA-DR beta chain is expressed higher in several types of immune cells with the RA-associated haplotype in comparison to RA-irrelevant haplotypes. By pre-selection of individuals with specific haplotypes in our study, we were able to decrease the level of heterogeneity and by accurate and haplotype-specific alignment to different MHC reference sequences, we were able to reliably identify levels of expression of these genes. Our results provide for the first time evidence for differential expression of several MHC Class II genes in whole blood, PBMCs, CD4+ T cells, CD8+ T cells, and CD14+ monocytes of individuals with predisposition to an autoimmune disease, i.e. RA.

The MHC is an extremely polymorphic region and quantification of expression of various allelic forms causes major issues for standard mapping methods and for studying expression of these genes. Using the standard human transcriptome reference, most of the sequencing reads will misalign to the MHC Class II locus. This could be recognized by for example *HLA-DRB5* expression within samples carrying *HLA-DRB1*\*04 alleles, where *HLA-DRB5* is not present (Figure 1). Here, we used the AltHapAlignR pipeline [8] for a more accurate and reliable expression analysis within the MHC region, which is especially important for the MHC Class II locus. It employs the eight available MHC reference haplotypes (APD, COX, DBB, MANN, MCF, PGF, QBL, and SSTO [22]) to generate less biased estimates of gene expression from this locus. However, the number of MHC reference haplotypes is still limited and obviously does not cover all patterns of the human genome. The standard MHC reference haplotype is PGF (*DRB1*\*15:01:01-*DQA1*\*01:02:01-*DQB1*\*06:02). The MHC reference haplotypes COX and QBL contain the *HLA-DRB1*\*03:01:01 allele (*DRB1*\*03:01:01-*DQA1*\*05:01:01-*DQB1*\*02:01:01), DBB and MANN contain the *HLA-DRB1*\*07:01:01 allele (*DRB1*\*07:01:01-*DQA1*\*02:01-*DQB1*\*03:03:02 or *DQB1*\*02:02, respectively), and SSTO contains the *HLA-DRB1*\*04:03:01 allele (*DRB1*\*04:03:01-*DQA1*\*03:01:01-*DQB1*\*03:05:01). In addition, *HLA-DRB1* is technically not present on the MHC reference haplotypes APD (*HLA-DRB1*\*13:01) and MCF (*HLA-DRB1*\*04:01 (GRCh38.p12)). Therefore, our findings showing that *HLA-DRB1* is lower expressed in *HLA-DRB1*\*13:01 individuals compared to *HLA-DRB1*\*04:01 and *15:01 are inconclusive. Furthermore, the *HLA-DRB1*\*04:01 samples are mapping to the closest matched MHC reference haplotype SSTO (*HLA-DRB1*\*04:03:01), which could potentially decrease efficiency of alignment in the MHC Class II locus towards a lower number of mapped sequencing reads. In this case, the detected level of *HLA-DRB1* expression for individuals with *HLA-DRB1*\*04:01 will be underestimated. The expression of *HLA-DRB4* gene, which is present at the *HLA-DRB1*\*04 haplotype and absent at *HLA-DRB1*\*03, *13, *15 haplotypes, was expected to be highly differentially expressed between individuals carrying *HLA-DRB1* SE-positive and SE-negative alleles. Even though there are several possible issues with the MHC reference haplotypes, our data shows that the expression of *HLA-DRB* and *HLA-DQ* genes is different between individuals carrying *HLA-DRB1*\*04:01 and *15:01 alleles in different cell types. Moreover, we found that *HLA-DRB1* tended to be higher expressed in *HLA-DRB1*\*04 and *07 alleles compared to *HLA-DRB1*\*03 and *15 alleles in both healthy individuals from the GTEx project and RA patients from EIRA/RECOMBINE study. Importantly, we studied the overall expression of *HLA-DRB1* and did not consider possible transcript heterogeneity due to alternative splicing that may cause the difference in expression.

To confirm the results of RNA-seq data, we performed qPCR analysis on the same PBMC and cell subset RNA samples. No primers could be designed to measure expression of the different forms of *HLA-DQA* (*HLA-DQA1* and *HLA-DQA2*) and *HLA-DQB* (*HLA-DQB1* and *HLA-DQB2*) due to gene homology. Therefore, we measured overall expression of *HLA-DQA* and *HLA-DQB*, and could confirm differences in expression by qPCR (Figure S1 and S2). In addition, no differences in expression of *HLA-DRA* could be detected by qPCR (Figure S3), which is also concordant with our RNA-seq data. No primers can be designed for simultaneous robust measure of expression of *HLA-DRB* in samples with different *HLA-DRB1* alleles and therefore the expression of *HLA-DRB* was not determined by qPCR in our study.

Using RNA-seq, we found *HLA-DRB1* and other MHC Class II genes to be differentially expressed in CD4+ T cells of healthy individuals carrying *HLA-DRB1* SE-positive versus SE-negative alleles. Although there are CD4+ T-cell subsets expressing HLA-DR [23, 24], by using xCell [20], a tool that performs cell type enrichment analysis from RNA-seq data, we noticed that suspensions of the CD4+ T cells in our experiment are slightly contaminated with monocytes (Figure S4). Monocytes express *HLA-DRB1* at a significantly higher level than CD4+ T cells and therefore we cannot totally exclude that the observed differences in the CD4+ T-cell subset are caused by the monocyte contamination.

HLA-DR is present on the surface of cells as heterodimers consisting of an alpha chain (HLA-DRA) and a beta chain (HLA-DRB1 together with either HLA-DRB3, HLA-DRB4 or HLA-DRB5, depending on the haplotype). Increased expression of HLA-DR is considered to be an activation marker on different cell types. Therefore, we correlated *HLA-DRB1* expression levels with the expression of other activation markers of different cell types (*CD25, CD38, CD69, CD86, CD40*, and *CD63*). We found no correlation with any of these activation markers (Figure S5-8), suggesting that the higher expression of *HLA-DRB1* in samples with *HLA-DRB1*\*04 alleles is not only a function of cell activation.

Molecular mechanisms of differences in gene expression levels for *HLA-DRB1* in immune cells of healthy individuals and RA patients depending on MHC haplotypes is not addressed in this study. Several studies have suggested that the levels of *HLA-DRB1* might be regulated by DNA methylation [27-29], which might be distinctive for different haplotypes. In addition, studies identifying functional consequences of higher gene expression levels of *HLA-DRB1* in *HLA-DRB1*\*04 individuals are also pending.

We attempted to confirm the differences in *HLA-DRB1* gene expression on protein level by western blotting. Several antibodies to detect HLA-DRB are currently on the market, however, no data on antibody binding to different allelic forms of HLA-DRB is available. Using isolated HLA-DRB1*03:01, HLA-DRB1*04:01, HLA-DRB1*15:01, and HLA-DRB5*01:01 proteins, we showed that the binding of the tested HLA-DRB antibodies to recombinant proteins was highly heterogeneous and haplotype specific (Figure 7A). Only two (ab133578 and DA2) out of the five tested antibodies can detect the recombinant protein HLA-DRB1*04:01. In addition, the binding of these antibodies is much stronger to HLA-DRB1*15:01 recombinant protein than to HLA-DRB1*04:01 and HLA-DRB1*03:01 recombinant proteins. Due to the differences in antibody binding to the different allelic forms of HLA-DRB, we generated a standard curve by using the recombinant proteins HLA-DRB1*04:01 and HLA-DRB1*15:01 as references in attempt to quantify the amount of HLA-DRB in the samples. However, in our experiments the expression of HLA-DRB is very variable in *HLA-DRB1*\*03:01/*04:01 samples, probably due to relatively weak binding of DA2 to HLA-DRB1*04:01. This prevented us from drawing a firm conclusion from these experiments about HLA-DRB protein levels. Further studies are needed to clarify if the beta chain of HLA-DR is differentially expressed on protein level in different cell types of healthy individuals and RA patients. It is worth mentioning that previous studies of expression of HLA-DR beta chain, either by western blot or by flow cytometry, should be taken with great caution due to uncontrolled extent of reactivities of commercial antibodies with polymorphic proteins (Figure 7A).

## 5. Conclusions

Using the available MHC reference haplotypes, we were able to identify by RNA-seq that *HLA-DRB* and *HLA-DQ* genes are differentially expressed in whole blood samples, PBMCs, CD4+ T cells, CD8+ T cells, and CD14+ monocytes of *HLA-DRB1* SE-positive versus SE-negative healthy individuals. In addition, we identified that *HLA-DRB1* is also differentially expressed in whole blood samples of *HLA-DRB1* SE-positive versus SE-negative RA patients. While MHC Class II genes consistently demonstrate differential expression between *HLA-DRB1* SE-positive and SE-negative healthy individuals, we found little difference for expression of non-MHC genes. In conclusion, our study shows that not only structural differences but also differences in expression of MHC Class II molecules in different immune cells may explain the relationships between immunity and autoimmune disease and presence of certain MHC Class II alleles in an individual.

## Data Availability

All sequencing data generated in this study are available at NCBI Gene Expression Omnibus accession number GSE163605.

https://www.ncbi.nlm.nih.gov/geo/query/acc.cgi?acc=GSE163605

## Author contributions

**Miranda Houtman:** Conceptualization, Methodology, Software, Validation, Formal analysis, Investigation, Data Curation, Writing – Original Draft, Visualization **Anna Dzebisashvili:** Investigation, Writing – Review & Editing **Espen Hesselberg:** Investigation, Writing – Review & Editing **Anatoly Dubnovitsky:** Resources, Writing – Review & Editing **Genadiy Kozhukh:** Resources, Writing – Review & Editing **Lars Rönnblom:** Resources, Writing – Review & Editing **Lars Klareskog:** Conceptualization, Writing – Review & Editing **Vivianne Malmström:** Writing – Review & Editing **Leonid Padyukov:** Conceptualization, Methodology, Data Curation, Writing – Review & Editing, Supervision, Project administration, Funding acquisition.

## Acknowledgements

We are very thankful to Dr. Yvonne Sundström, Dr. Danika Schepis, and Dr. Louise Berg for running flow cytometry on PBMCs and sorted cell populations, and Dr. Karolina Tandre and Dr. Maija-Leena Eloranta for sample collection. Transcriptomic profiling was performed by the SNP&SEQ Technology Platform in Uppsala. This facility is part of the National Genomics Infrastructure (NGI) Sweden and Science for Life Laboratory. The SNP&SEQ Technology is also supported by the Swedish Research Council and the Knut and Alice Wallenberg Foundation. The computations were performed on resources provided by SNIC through Uppsala Multidisciplinary Centre for Advanced Computational Science (UPPMAX), supported by NGI Sweden. This work was supported by the Swedish Research Council [2015-3006, 2018-2399 and 2018-2884], the Swedish Rheumatism Association, King Gustav V’s 80-year Foundation and Börje Dahlin Foundation.

