## Supplementary material for "Expression levels of *HLA-DRB* and *HLA-DQ* are associated with MHC Class II haplotypes in healthy individuals and rheumatoid arthritis patients": Figures S1-S8

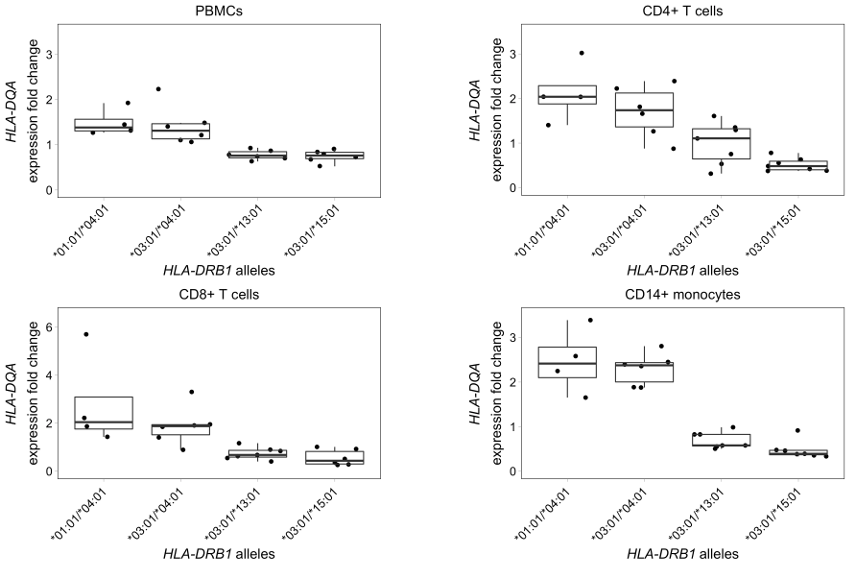

**Figure S1. *HLA-DQA* expression levels in different cell types of healthy individuals carrying different *HLA-DRB1* alleles.** *HLA-DQA* expression in PBMCs, CD4+ and CD8+ T cells, and CD14+ monocytes of individuals carrying *HLA-DRB1**01:01/*04:01, *HLA-DRB1**03:01/*04:01, *HLA-DRB1**03:01/13:01, and *HLA-DRB1**03:01/*15:01 alleles obtained by quantitative real-time PCR. The endogenous controls *ACTIN*, *UBE2D2* and *ZNF592* were used to normalize the expression levels of *HLA-DQA*.

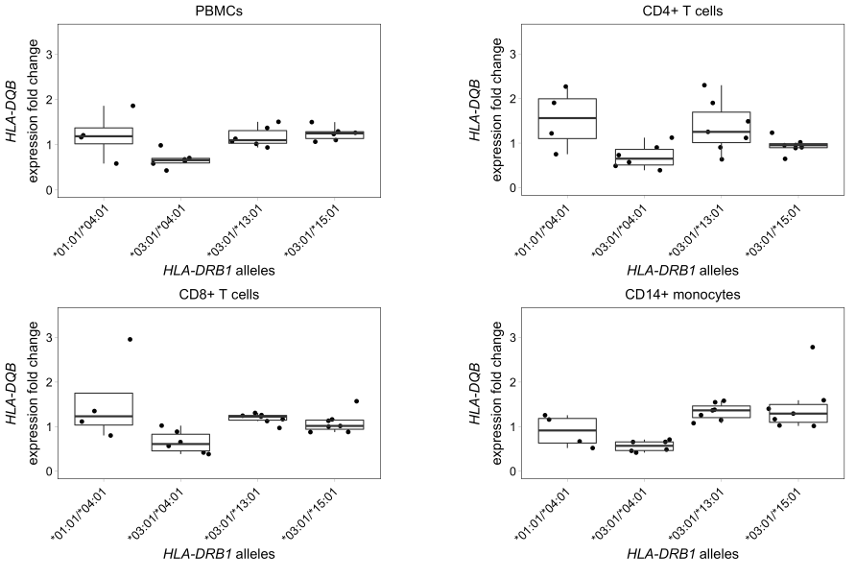

**Figure S2. *HLA-DQB* expression levels in different cell types of healthy individuals carrying different *HLA-DRB1* alleles.** *HLA-DQB* expression in PBMCs, CD4+ and CD8+ T cells, and CD14+ monocytes of individuals carrying *HLA-DRB1**01:01/*04:01, *HLA-DRB1**03:01/*04:01, *HLA-DRB1**03:01/13:01, and *HLA-DRB1**03:01/*15:01 alleles obtained by quantitative real-time PCR. The endogenous controls *ACTIN*, *UBE2D2* and *ZNF592* were used to normalize the expression levels of *HLA-DQB*.

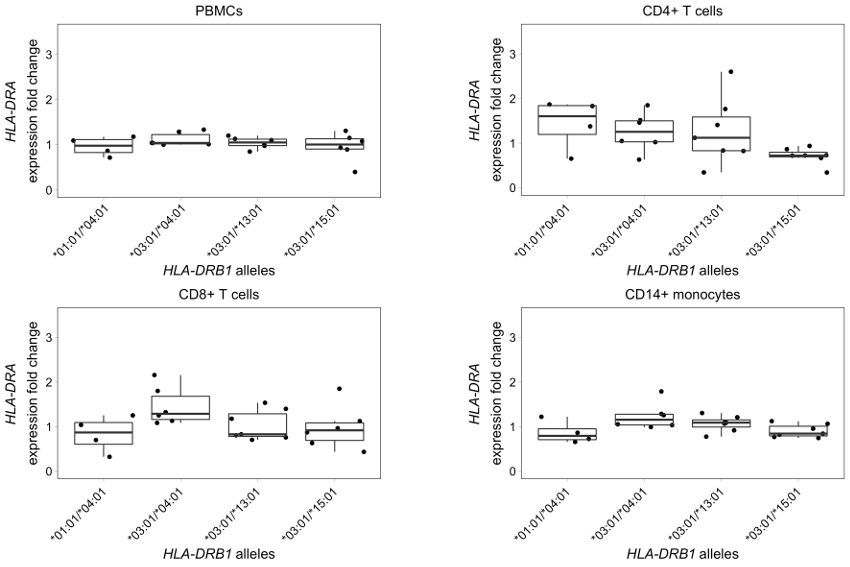

**Figure S3. *HLA-DRA* expression levels in different cell types of healthy individuals carrying different *HLA-DRB1* alleles.** *HLA-DRA* expression in PBMCs, CD4+ and CD8+ T cells, and CD14+ monocytes of individuals carrying *HLA-DRB1**01:01/*04:01, *HLA-DRB1**03:01/*04:01, *HLA-DRB1**03:01/13:01, and *HLA-DRB1**03:01/*15:01 alleles obtained by quantitative real-time PCR. The endogenous controls *ACTIN*, *UBE2D2* and *ZNF592* were used to normalize the expression levels of *HLA-DRA*.

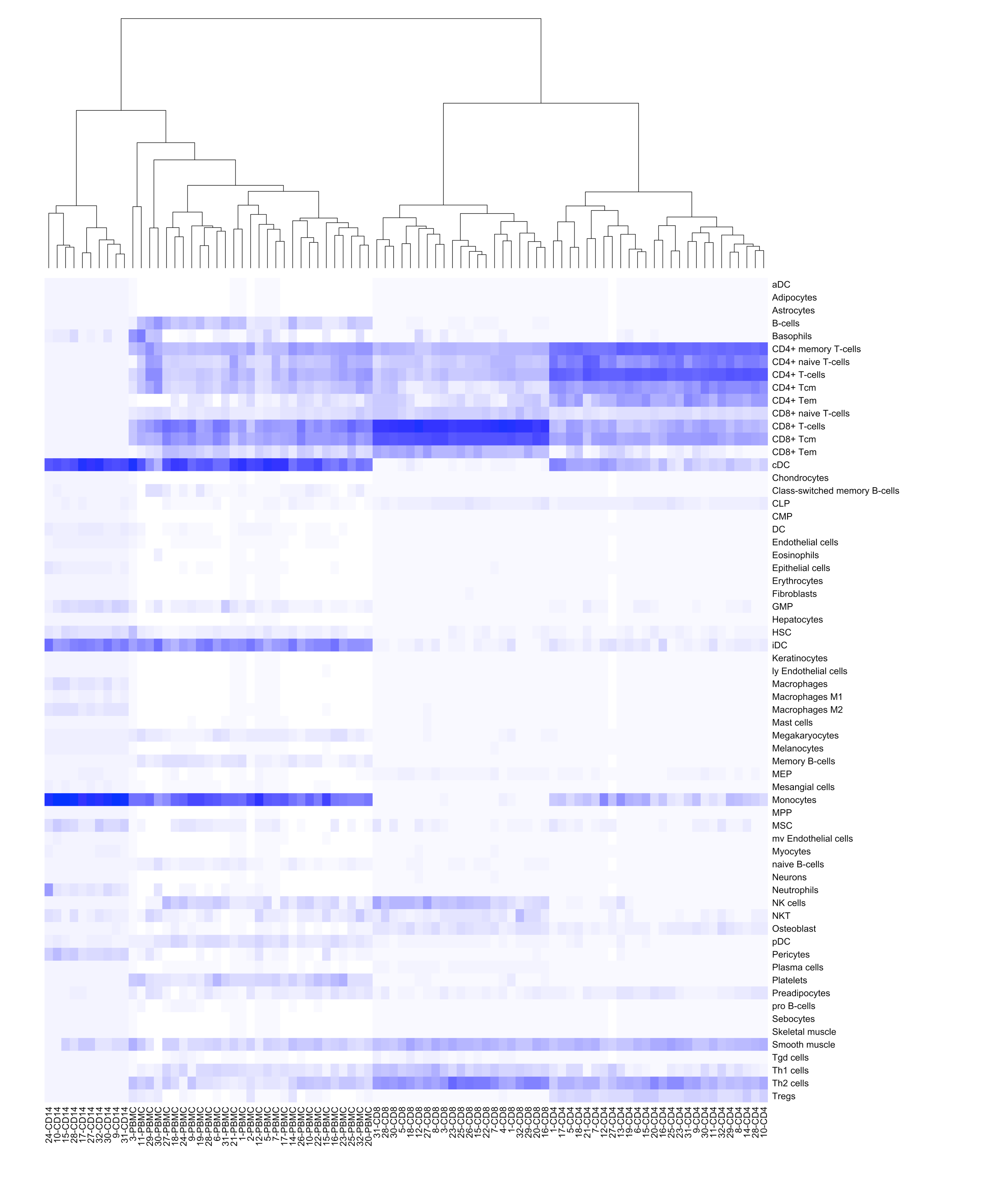

**Figure S4. Cell type enrichment analysis in the different isolated cell subsets.** Clustering heatmap showing cellular heterogeneity in PBMC, CD4+ T-cell, CD8+ T-cell, and CD14+ monocyte subsets. The xCell tool [13] was used to identify cellular heterogeneity in the different isolated cell subsets from gene expression data. Intensity of the blue color indicates relative cell type enrichment.

**Figure S5. Correlation of gene expression levels of *HLA-DRB1* and other activation markers in PBMCs of healthy individuals.** Expression levels of *HLA-DRB1*, *CD25*, *CD38*, *CD69*, *CD86*, *CD40*, and *CD63* are shown as log transformed normalized gene counts. In each subplot, cor is Pearson correlation.

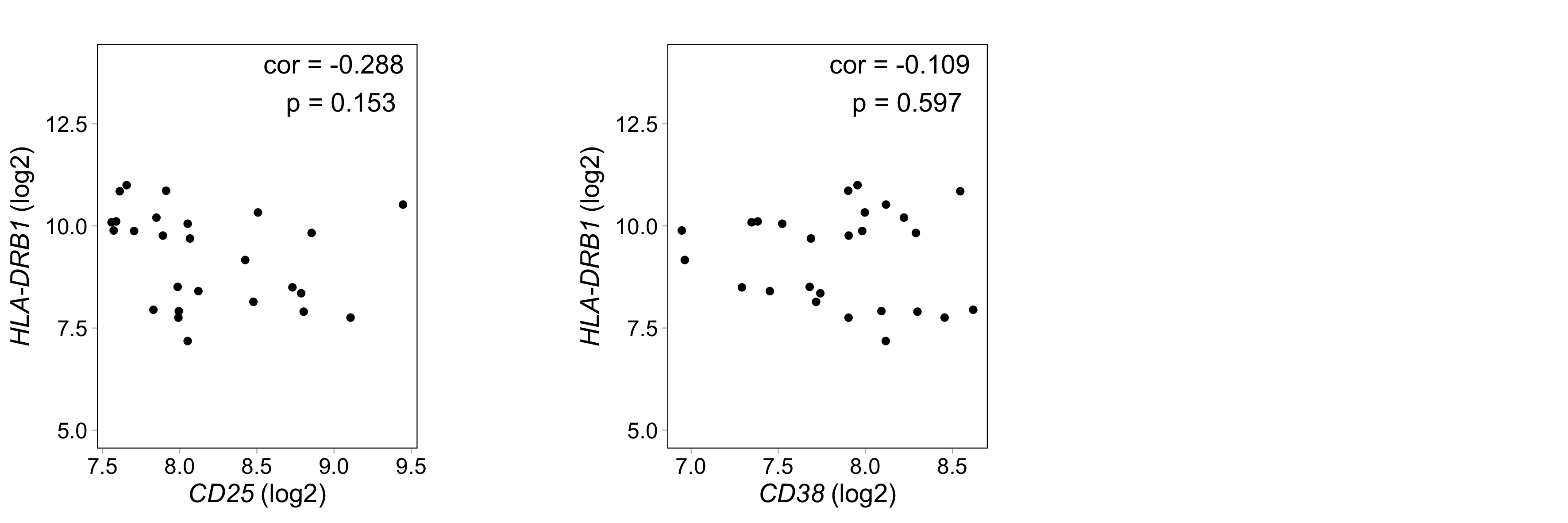

**Figure S6. Correlation of gene expression levels of *HLA-DRB1* and other activation markers in CD4+ T cells of healthy individuals.** Expression levels of *HLA-DRB1*, *CD25*, and *CD38* are shown as log transformed normalized gene counts. In each subplot, cor is Pearson correlation.

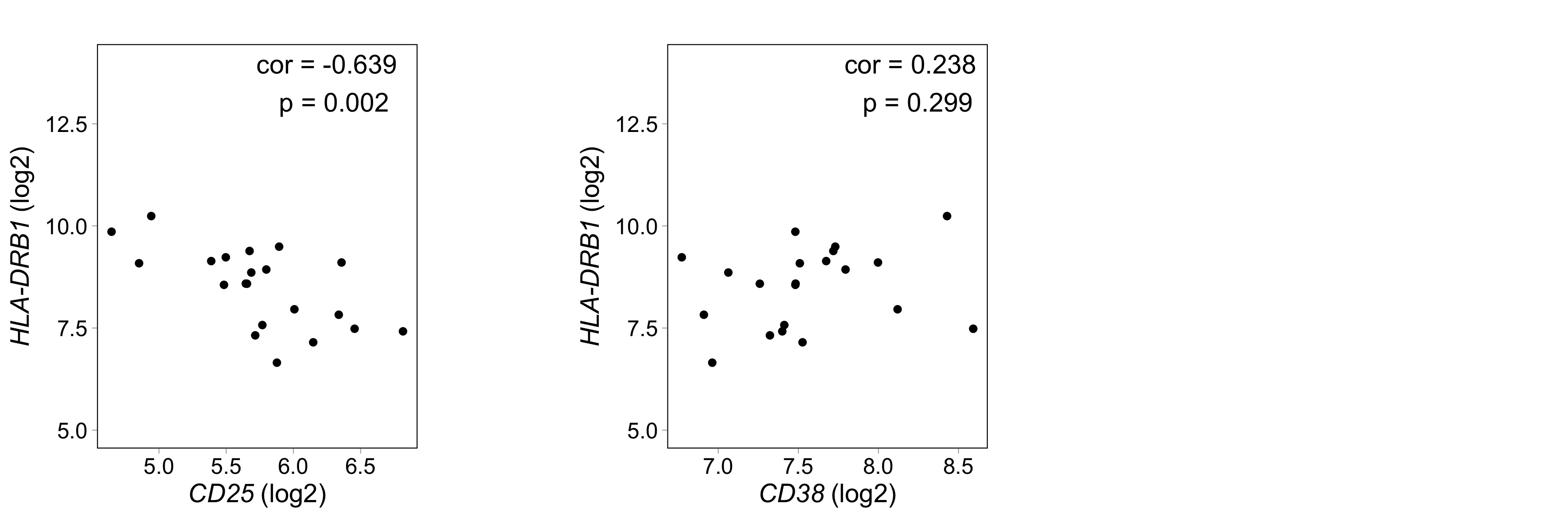

**Figure S7. Correlation of gene expression levels of *HLA-DRB1* and other activation markers in CD8+ T cells of healthy individuals.** Expression levels of *HLA-DRB1*, *CD25*, and *CD38* are shown as log transformed normalized gene counts. In each subplot, cor is Pearson correlation.

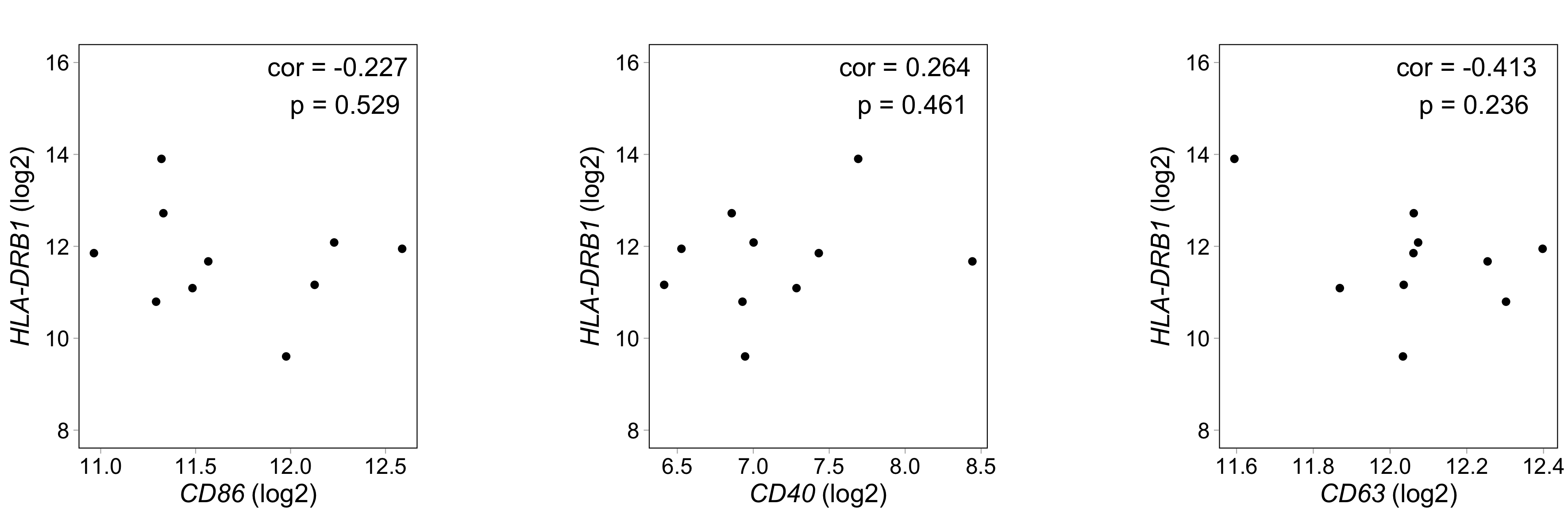

**Figure S8. Correlation of gene expression levels of *HLA-DRB1* and other activation markers in CD14+ monocytes of healthy individuals.** Expression levels of *HLA-DRB1*, *CD86*, *CD40*, and *CD63* are shown as log transformed normalized gene counts. In each subplot, cor is Pearson correlation.
